# Sexual abuse and harsh punishment in early life are associated with more obsessive-compulsive symptoms in adulthood: An online survey

**DOI:** 10.1101/2022.11.03.22281919

**Authors:** M. Król, Y. Cao, E.J. Kirkham

## Abstract

**Introduction:** Childhood and adolescent maltreatment (CAM) is associated with many psychiatric conditions, including obsessive-compulsive disorder (OCD). However, it remains unclear whether the severity of OCD symptoms vary according to the type of stress encountered in early life.

**Method:** A sample of 345 participants (174 of whom had OCD) completed online measures of CAM (Child Abuse and Trauma Scale; CATS), OCD (Obsessive-Compulsive Inventory-Revised; OCI-R), anxiety, and depression (Hospital Anxiety and Depression Scale; HADS).

Regression analysis was used to examine associations between OCD symptoms and four subtypes of CAM: emotional abuse, neglect/home environment, punishment, and sexual abuse. Anxiety and depression were included as control variables.

**Results:** Higher levels of sexual abuse and punishment were significantly associated with more OCD symptoms irrespective of scores on measures of anxiety and depression. Emotional abuse and neglect/home environment were not significantly associated with OCD symptoms.

**Discussion:** These findings highlight the prospect of a specific role for harsh punishment and sexual abuse in the development of OCD. Future studies should examine this possibility using longitudinal designs. Health professionals should be mindful that individuals with OCD may have experienced heightened levels of CAM, especially in the domains of punishment and sexual abuse.

## Introduction

Early life stress (ELS) refers to a broad range of adverse experiences, such as abuse or parental loss, which occur during childhood or adolescence. Within the field of ELS, attention has been drawn to an individual’s experience in their home environment, with a focus on experiences of abuse and neglect, referred to henceforth as child and adolescent maltreatment (CAM). CAM has been associated with a range of mental health conditions in adults (Bowen et al., 2018; Heim & Nemeroff, 2001; Krupnick et al., 2004; Norman et al., 2012), including post-traumatic stress disorder (PTSD; Widom, 1999), schizophrenia (Read et al., 2005), anxiety (Heim & Nemeroff, 2001), and depression (Nanni, Uher & Danese, 2012). Though the relationship between CAM and mental illness is well-established, it remains unclear whether the type of stress encountered in early life is related to the form or severity of mental health condition(s) experienced in adulthood.

Obsessive-compulsive disorder (OCD) has also been associated with CAM (ref), though compared to other health conditions it has received relatively little attention in the CAM literature (Destrée et al., 2021). OCD is prevalent among the general population (2.5% – 3%) and has a substantial health and economic cost to society (Robbins, Vaghi & Banca, 2019). It is characterised by recurrent intrusive and unwanted thoughts, urges or images (obsessions) and behaviours or mental acts performed repetitively (compulsions; Veale & Roberts, 2014). Both obsessions and compulsions are associated with high levels of distress and anxiety. OCD is often chronic (Tibi et al., 2020), and responsible for high levels of impairment in daily life, affecting relationships and social functioning (Ruscio et al., 2010). Understanding the risk factors associated with OCD is therefore an essential task for psychological research.

Previous studies suggest that people with OCD who experienced high levels of CAM may show more severe symptoms (Gershuny et al., 2008) and have a reduced response to treatment (Dykshoorn, 2014). Regarding subtypes of CAM, a number of studies have highlighted relationships between OCD and early emotional abuse (Mathews et al., 2008; Ou et al., 2021; Hemmings et al., 2013), and between OCD and early emotional or physical neglect (Lochner et al., 2002; Hemmings et al., 2013; Mathews et al., 2008; Brooks et al., 2016). A smaller number of studies have also reported worse OCD outcomes in people who experienced physical or sexual abuse in early life (Gershuny et al., 2008; Ou et al., 2021). However, a thorough understanding of the relationships between specific types of CAM and OCD symptoms in adulthood remains elusive.

Both CAM and OCD are positively associated with anxiety and depression (Sun et al., 2015). However, there is conflicting evidence as to whether the relationship between CAM and OCD is direct or mediated by comorbid anxiety and/or depression. Mathews et al. (2008) found that the relationship between CAM and OCD was independent of comorbid anxiety, while other studies have found that the relationship between CAM and OCD is fully mediated by anxiety and depression (Briggs & Price, 2009; Çoban & Tan, 2020). In light of this, it is essential that any examination of the relationship between CAM and OCD accounts for comorbid anxiety and depression.

Therefore the present study sought to examine the relationship between CAM subtypes and OCD symptoms, while controlling for comorbid anxiety and depression. A sample of adults with and without OCD took part in an online survey which included measures of CAM (emotional abuse, sexual abuse, punishment and neglect), OCD, anxiety and depression. It was predicted that OCD would be associated with multiple subtypes of CAM, and that these relationships would persist after controlling for anxiety and depression.

## Method

### Participants

Recruitment elicited responses from 392 participants. Of these, 47 were excluded from the present analyses due to missing or unreliable (*n* = 2) data, resulting in a sample size of 345. In most cases, missing data was due to the participant choosing not to view the early life stress questionnaire, an option which was added to the survey during the ethical review process. People with OCD were deliberately over-sampled to ensure variability in the measure of OCD and to ensure that any findings would be based on and applicable to people with OCD. The number of participants who reported that they had OCD was 174 (50.4%), with 78 of these reporting that they had been given a clinical diagnosis. The number of participants who met criteria for OCD according to the recommended cut-off on the OCI-R measure (>= 21) was 194 (56.2%). The mean age was 29.90 (*SD* = 10.34). Demographic characteristics of participants are detailed in Table 1.

**Table 1.** Demographic characteristics of participants.

|  | Variable | *n* | Percentage |
| --- | --- | --- | --- |
| Gender |  |  |  |
|  | Female | 226 | 65.7% |
|  | Male | 111 | 32.3% |
|  | Other | 7 | 2.0% |
| Ethnicity |  |  |  |
|  | Asian or Asian British | 21 | 6.1% |
|  | Black, Black British, Caribbean, or African | 33 | 9.6% |
|  | Mixed or multiple ethnic groups | 19 | 5.5% |
|  | White | 258 | 74.8% |
|  | Other ethnic group | 14 | 4.1% |
| Attended university |  |  |  |
|  | Yes | 297 | 86.1% |
|  | No | 48 | 13.9% |

The project received ethical approval from the Philosophy, Psychology and Language Sciences Research Ethics Committee at the University of Edinburgh (Reference Number: 227-2122/11). All participants completed an online informed consent form and were made aware that they could withdraw from the study at any time. Approximately 200 participants took part through Prolific Academic (www.prolific.ac) and were paid an average of £2.25 upon completion of the survey. Prolific Academic allows researchers to advertise their survey to specific groups of people based on pre-screening criteria held within the Prolific Academic environment. This allows for the selection of people who have a mental health condition.

However, there is no pre-screening question for OCD specifically. Therefore, a short survey was conducted that asked Prolific users who had a mental health condition whether they had experienced obsessive-compulsive symptoms. Participants who stated that they had were then invited to take part in the full survey. The rest of the participants were recruited through social media platforms and survey exchange communities (www.surveycircle.com; www.surveyswap.io). These participants were asked to choose one of two UK OCD charities (OCD UK or OCD Action) to receive a £4 donation.

### Materials

The Obsessive-Compulsive Inventory-Revised (OCI-R; Foa et al., 2002) was used to measure the severity of participants’ obsessive-compulsive symptoms. It consists of 18 items (e.g. “I check things more often than necessary”, “I am upset by unpleasant thoughts that come into my mind against my will”) measured on a 5-item Likert scale ranging from 0 (Not at all) to 4 (Extremely). Possible scores range from 0 to 72, and scores at or above 21 indicate the likely presence of OCD. The OCI-R has been found to be psychometrically sound and appropriate for clinical and non-clinical populations and research purposes (Hajcak et al., 2004; Huppert et al., 2007). The scale showed excellent internal consistency within the current sample (*n* = 345, *α* = 0.93).

The Child Abuse and Trauma Scale (CATS; Sanders & Becker-Lausen, 1995) was used to measure the frequency and extent of different types of childhood and adolescence maltreatment. It is a 38-item questionnaire measured on a 5-point Likert scale ranging from 0 (Never) to 4 (Always). CATS scores are calculated as mean scores, resulting in a score between 0 and 4 for the full CATS measure and each of the subscales. Higher scores indicate a greater frequency and severity of childhood and adolescence abuse and traumatic experiences. The CATS consists of four subscales: negative home environment/neglect, punishment, sexual abuse (Sanders & Becker-Lausen, 1995), and emotional abuse (Kent & Waller, 1998). Example items include “As a child, did you feel unwanted or emotionally neglected?”; “Were there traumatic or upsetting sexual experiences when you were a child or teenager that you couldn’t speak to adults about?”; and “Did your parents ever hit or beat you when you did not expect it?”. The CATS has shown good test-retest reliability and validity (Kent and Waller, 1998; Sanders & Becker-Lausen, 1995). The scale showed excellent internal consistency within the current sample (*n* = 345, *α* = 0.95).

The Hospital Anxiety and Depression Scale (HADS; Zigmond & Snaith,1983) was used to measure respondents’ levels of anxiety and depression. HADS is a 14-item self-report measure with response options 0 to 3 for each question. It provides a measure of anxiety and a measure of depression, each between 0 and 21. Scores greater than or equal to 11 indicate severe levels of anxiety/depression. An example item for the HADS-anxiety is “I get sudden feelings of panic”; an example item for HADS-depression is “I still enjoy the things I used to enjoy”. HADS is regarded as a valid measure among the general population as well as psychiatric and primary care patients (Bjelland et al., 2002). Good internal consistency was shown within the current sample for both the anxiety subscale (*n* = 345, *α* = 0.83) and the depression subscale (*n* = 345, *α* = 0.83).

### Statistical analysis

In cases where all values on a given questionnaire were missing, the case was removed from analyses. A number of cases (*n* = 64) had missing data for one or more individual items on the CATS. Therefore, multiple imputation was used to impute values for these items. In each case, five imputations were generated and the mean of these five imputations was used as the final imputed value. For the OCI-R, there were two cases in which one item value was missing. In these two cases the mean of the remaining values for the relevant subscale was entered in place of the missing value.

Hierarchical multiple regression was used to examine which CATS subscales (emotional abuse, neglect/negative home environment, punishment and sexual abuse) were independently associated with OCD scores. The control variables of anxiety and depression were entered in Model 1. In Model 2, the four CATS subscales were entered in addition to anxiety and depression (Table 3).

## Results

Mean scores on each variable of interest are reported in Table 2. Participants with self-reported OCD had significantly higher scores than people without self-reported OCD on questionnaire measures of current OCD symptoms, and on measures of child and adolescent maltreatment overall, emotional abuse, neglect, and punishment. There were no significant differences between the two groups on experience of sexual abuse, nor on current anxiety or depression.

**Table 2.**
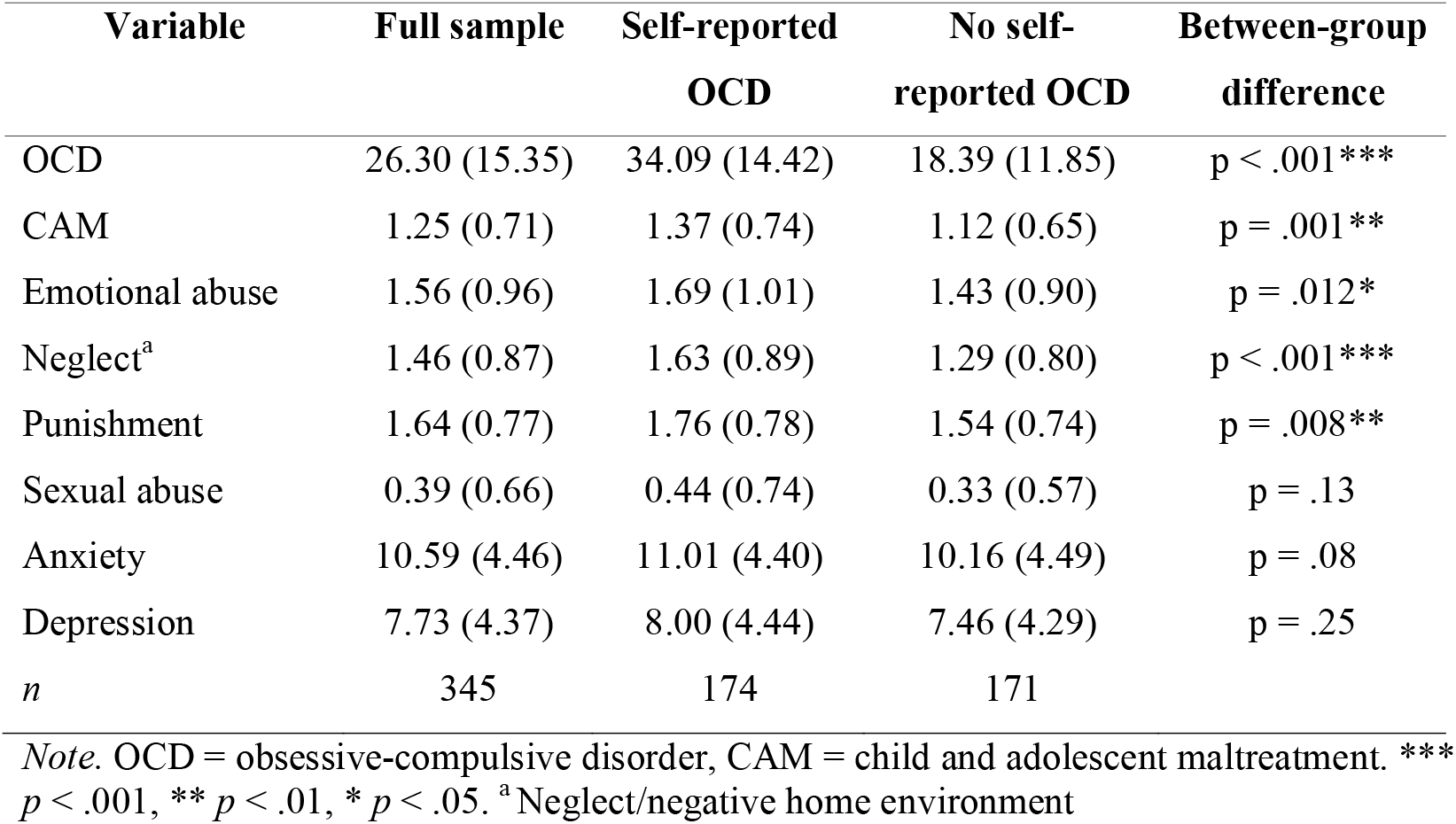
Mean scores and standard deviations on variables of interest by OCD group

Hierarchical multiple regression was used to examine which CATS subscales (neglect/negative home environment, punishment, sexual abuse and emotional abuse) were independently associated with OCD scores (Table 3). One case had a standard residual greater than 3, though this case did not have a high leverage value or Cook’s distance and therefore was not removed from the analysis. The full model (Model 2) of anxiety, depression, neglect/negative home environment, punishment, sexual abuse and emotional abuse was significant, *R*^2^ = .20, *F*(6, 338) = 13.70, *p* < .001; adjusted *R*^2^ = .18. The addition of the CATS subscales to the model (Model 2) explained a significant additional amount of variance in OCD scores, *R*^2^ change = .18, *F*(4, 338) = 18.90, *p* < .001.

**Table 3.** Association between types of ELS and OCD symptoms when controlling for anxiety and depression

| Variable | OCD score |  |  |  |  |  |
| --- | --- | --- | --- | --- | --- | --- |
|  | Model 1 |  |  | Model 2 |  |  |
| | $\beta$ | $B$ | 95% CI | $\beta$ | $B$ | 95% CI |
| Anxiety | 0.12 | 0.43 | -0.03, 0.88 | 0.08 | 0.27 | -0.15, 0.69 |
| Depression | -0.15 | -0.54* | -1.01, -0.07 | -0.12 | -0.41 | -0.84, 0.02 |
| Emotional abuse |  |  |  | 0.05 | 0.77 | -2.28, 3.82 |
| Neglect<sup>a</sup> |  |  |  | 0.17 | 3.07 | -0.19, 6.33 |
| Punishment |  |  |  | 0.16 | 3.20* | 0.57, 5.83 |
| Sexual abuse |  |  |  | 0.14 | 3.28* | 0.63, 5.93 |
| R<sup>2</sup> | 0.02 |  |  | 0.20*** |  |  |
| $\Delta R^2$ | 0.02 | | | 0.18*** | | |

Note. \* $p < .05$ , \*\*\* $p < .001$ , $\Delta$ = change, <sup>a</sup>Neglect/negative home environment

## Discussion

This study investigated the relationship between different subtypes of childhood and adolescent maltreatment and obsessive-compulsive symptoms in adulthood in a sample of participants with and without OCD. It was found that, when controlling for anxiety and depression, more obsessive-compulsive symptoms were associated with higher scores on measures of two CAM subtypes: punishment and sexual abuse. OCD symptoms were not significantly associated with the CAM subtypes of neglect or emotional abuse.

These findings are partly supported by previous literature (McGregor et al., 2016; Lochner et al., 2004). Bey et al. (2017) found that participants with OCD had experienced significantly more childhood trauma than unaffected first-degree relatives, and that specific OCD symptoms (obsessions/checking) were related to more experience of early sexual abuse. Similarly, Renkema et al. (2020) reported a significant relationship between sexual abuse and obsessive-compulsive symptoms in a sample of patients with psychosis and their siblings. Moreover, a case-control study found that among all types of CAM, only sexual abuse was related to suicidal ideation in patients with OCD (Khosravani et al., 2017).

The relationship between the CAM subtype of punishment and obsessive-compulsive symptoms is particularly noteworthy, as it has not been reported as frequently in the prior literature. This may be due to methodological factors; specifically, most studies investigating childhood trauma and OCD use the Childhood Trauma Questionnaire (CTQ; Bernstein & Fink, 1998), whereas the present study used the CATS. While the two scales seek to measure the same overarching construct of CAM, there are some differences in the way specific subtypes are classified and measured. The CTQ refers to punishment in only one of its questions, within the physical abuse subscale. This physical abuse subscale mainly covers experiences of severe physical harm. In contrast, most CATS questions regarding punishment did not directly imply physical harm. Rather, the construct of “punishment” in the CATS encompasses both physical abuse and broader conceptualisations of punishment, with a focus on adherence to strict house rules.

In light of this, the finding of a relationship between the CATS punishment subscale and OCD symptoms may be best understood in reference to literature on parenting styles and OCD. Being brought up under an authoritarian parenting style, which involves strict adherence to rules and low levels of nurturing, has been associated with more OCD symptoms later in life (Timpano et al., 2010). Future research should examine whether a measure of parenting style would explain variance in OCD symptoms over and above the measure of punishment in the CATS.

Whilst the finding of positive relationships between CAM and adult OCD symptoms is in line with much of the previous literature (refs), it was surprising that the present study did not find evidence of a relationship between emotional abuse or neglect and OCD symptoms. One speculative explanation for this finding is that most of the studies that found emotional abuse to be linked to obsessive-compulsive symptoms were conducted in outpatient settings (Bey et al., 2017; Hemmings et al., 2013; Renkema et al., 2020). In contrast, participants in the present research were recruited from the general population using online self-report measures. Other studies with comparable participant samples have not focused on CAM subscales but rather on the overall effect of CAM on obsessive-compulsive symptoms (Briggs & Price, 2009; Kroska et al., 2018). As discussed previously, the use of the CATS to measure CAM may also explain discrepancies between the present study and previous research. Notably, the “neglect” subscale in the CATS covers emotional neglect, physical neglect and “negative home environment” (e.g. parental substance misuse or fighting). It is therefore unlikely to assess the same construct as the other, more specific measures of neglect used in prior research (e.g. Lochner et al. 2004; Hemmings et al. 2013; Carpenter & Chung, 2011).

A key finding of the present research was that the positive relationships between subtypes of CAM and OCD symptoms remained significant after controlling for depression and anxiety. This is noteworthy given that CAM is associated with most mental health conditions, and previous research has been equivocal on whether there are specific relationships between CAM and OCD which are not explained by comorbid anxiety (Çoban & Tan, 2020; Briggs & Price, 2009; Mathews et al., 2008). The present finding of a relationship between OCD and the CAM subtypes of punishment and sexual abuse offers a promising direction for future work which seeks to understand why an individual who experiences CAM will develop OCD rather than a different mental health condition.

To the authors’ knowledge, this is the first examination of relationships between subtypes of CAM and OCD in a sample recruited from the general population (as opposed to from health-related settings). Furthermore, the ability to examine a wide range of obsessive-compulsive symptoms was facilitated by the recruitment of a sample in which approximately half of respondents had OCD and half did not. Nevertheless, the study was subject to a number of limitations. Firstly, the retrospective nature of the CATS leaves it open to bias and under-reporting of CAM (Widom et al., 2004). Nonetheless, a review of the evidence of validity of retrospective reports of adverse childhood experience showed that instances when CAM is reported to have occurred are generally correct (Hardt & Rutter, 2004). Secondly, the use of the CATS scale for assessing CAM hinders comparisons with other studies, which have typically used the CTQ. Thirdly, as the study was completed online unsupervised, there may be some inaccuracy in the data, and self-reported clinical diagnoses of OCD could not be verified. Finally, the data were cross-sectional. This places limits on the extent to which causality could be inferred.

In light of this, future research should collect prospective longitudinal data, or draw upon existing cohort studies. Such an approach would allow for a greater understanding of causal relationships and would reduce recall bias. It would also allow for investigation of additional research questions, such as whether experience of CAM is related to the age of onset of OCD, and whether OCD which occurs following CAM emerges before or after comorbid mental health conditions.

To summarise, this study provides further evidence for a relationship between maltreatment in childhood and adolescence and OCD symptoms in adulthood. It was found that early life sexual abuse and punishment are specifically related to obsessive-compulsive symptoms, and that this relationship is independent of comorbid anxiety and depression. Professionals who work with people with OCD should be mindful that this population is likely to have experienced elevated levels of CAM, which may have been particularly apparent in the domains of sexual abuse and harsh parental punishment.

## Data Availability

De-identified data produced in the present study are available upon reasonable request to the authors.

